# Evaluation of Acute Myeloid Leukemia Genomes using Genomic Proximity Mapping

**DOI:** 10.1101/2024.05.31.24308228

**Authors:** Cecilia CS Yeung, Stephen M. Eacker, Olga Sala-Torra, Lan Beppu, David W. Woolston, Ivan Liachko, Maika Malig, Derek Stirewalt, Min Fang, Jerald Radich

**Affiliations:** Translational Science and Transplantation Division, Fred Hutchinson Cancer Center, Seattle, WA, USA; Phase Genomics, Seattle, WA, USA; Department of Medicine, University of Washington Medical Center, Seattle, WA, USA; Department of Laboratory Medicine and Pathology, University of Washington Medical Center, Seattle, WA, USA

**Keywords:** Acute myeloid leukemia, AML, genomic proximity mapping, molecular genetics pathology, genomic profiling

## Abstract

**Background:** Cytogenetic analysis encompasses a suite of standard-of-care diagnostic testing methods that is routinely applied in cases of acute myeloid leukemia (AML) to assess chromosomal changes that are clinically relevant for risk classification and treatment decisions.

**Objective:** In this study, we assess the use of Genomic Proximity Mapping (GPM) for cytogenomic analysis of AML diagnostic specimens for detection of cytogenetic risk variants included in the European Leukemia Network (ELN) risk stratification guidelines.

**Methods:** Archival patient samples (N=48) from the Fred Hutchinson Cancer Center leukemia bank with historical clinical cytogenetic data were processed for GPM and analyzed with the CytoTerra® cloud-based analysis platform.

**Results:** GPM showed 100% concordance for all specific variants that have associated impacts on risk stratification as defined by ELN 2022 criteria, and a 72% concordance rate when considering all variants reported by the FH cytogenetic lab. GPM identified 39 additional variants, including variants of known clinical impact, not observed by cytogenetics.

**Conclusions:** GPM is an effective solution for the evaluation of known AML-associated risk variants and a source for biomarker discovery.

## Introduction

Cytogenetic analysis encompasses a suite of standard-of-care diagnostic testing that is routinely applied in cases of acute myeloid leukemia (AML) and other hematological malignancies(1). Decades of studies using karyotyping, fluorescence in situ hybridization (FISH), and, more recently, chromosomal microarrays (CMA) have identified recurrent translocations, inversion, deletions, duplications, and copy-neutral loss of heterozygosity in AML. These recurrent chromosome aberrations result in the generation of oncogenic fusion genes, disrupt tumor suppressor genes, or amplify oncogenes; information that can be used to assess patient risk and guide treatment decisions(2). Cytogenetic testing methods each have a non-overlapping set of strengths, including genome-wide variant detection (CMA and karyotyping), target specificity (FISH and CMA), and ability to detect balanced rearrangements (FISH and karyotyping). There are also technical challenges and limitations presented by these standard methods, including low resolution (karyotyping and FISH), the need for live, dividing cells (karyotyping), limited scope of detection (FISH), and inability to detect balanced rearrangements (CMA). These partially overlapping strengths and limitations often necessitate multi-modality testing for AML diagnosis and risk stratification. Nonetheless, cytogenetic assessments are important measures of the risk classification schemes used in AML treatment and clinical trial design.

We have developed a next-generation approach for the identification of chromosome aberrations to complement and improve testing using multiple modalities that are required to meet diagnostic testing needs in AML. This method uses Genomic Proximity Mapping™ (GPM), a next-generation sequencing (NGS)-based assay that uses the proximity of interacting DNA sequences in intact cells to determine the linear structure of chromosomes(3, 4). The method involves crosslinking intact nuclei, freezing chromatin interactions in place (Fig. 1A). Crosslinked chromatin is fragmented and chimeric DNA molecules are generated through the ligation of DNA molecules that were in close three-dimensional proximity in the nucleus(5). Sequences that are closer together along the linear length of a chromosome are more likely to be proximal and interact with each other than sequences that are distant (Fig. 1B). Using these data, it is possible to identify structural variants (SVs) by detecting the changes in pairwise interaction frequencies across the genome. The frequency of pairwise sequence interactions can be represented as a heatmap (Fig. 1C). In normal genomes, interaction signal is primarily limited to interactions within a chromosome. However, in leukemia genomes, interchromosomal SVs such as translocations are commonly observed. These chromosome rearrangements are easily visualized as deviations from the expected pattern of pairwise interactions on the heatmap. The pattern of deviation from expected interactions can be used to interpret the structure of both inter- and intrachromosomal SVs including translocations, inversions, insertions, deletions and duplications (Fig. 1D).

**Fig. 1.**
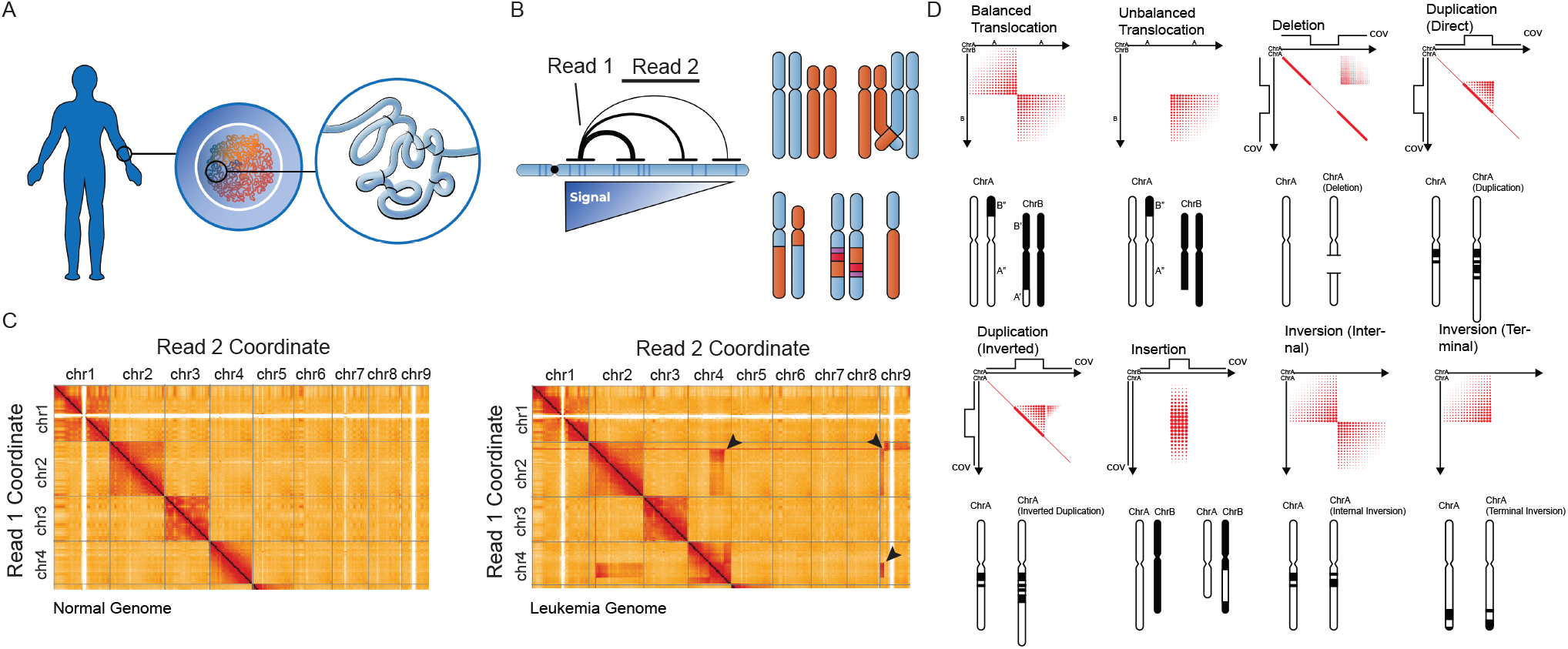
Principles of genomic proximity mapping (GPM). (A) Cellular samples are collected from patients and subjected to crosslinking while still intact, freezing native chromatin conformation in place prior to proximity ligation and library generation. (B) The frequency that pairs of sequences physically interact is governed primarily by their distance along the linear length of a chromosome. Using this information, the CytoTerra variant callers can identify abnormalities in chromosome structure. (C) A view of interaction heatmap generated for a normal (left) and leukemic genome (right). Arrowheads indicate deviation from expected patterns of interaction, in this example highlighting translocations. (D) A visual guide to how classes of chromosome aberrations appear on the GPM sequence interaction matrix. Genomic coordinates are mirrored on X and Y axes while sequence interaction frequency is represented with increasing intensity on the heat map. Using a combination of interaction frequency and sequencing coverage depth, GPM can identify every major class of structural variation.

In this study we assess the use of GPM platform CytoTerra® as a method for cytogenomic analysis for AML. Using cryopreserved diagnostic specimens, we show that CytoTerra detects all known risk variants included in the European Leukemia Network (ELN) risk stratification guidelines(2) that were previously identified by standard-of-care cytogenetics. However, GPM analysis identified additional variants of known clinical importance that were absent from cytogenetic reports at the time of diagnosis. In the study population of 48 samples, we identified a recurrent inversion not previously observed in AML. These observations support the effectiveness of GPM to enhance cytogenomic characterization of AML cases.

## Materials and methods

### A. Study population and clinical cytogenetic evaluation

The experiments were undertaken with the understanding and written consent of each subject, and that the study conforms with The Code of Ethics of the World Medical Association (Declaration of Helsinki), printed in the British Medical Journal (18 July 1964). All patients were consented with the approval of Fred Hutchinson Cancer Center (FH) Institutional Review Board. Inclusion criteria includes a confirmed diagnosis of AML, specific consent for whole genome sequencing, and multiple cryopreserved vials in the FH leukemia bank. Samples were specifically chosen to include a variety of different AML-associated genetic abnormalities identified by standard-of-care testing methods at FH. Data from institutional medical records were obtained and verified manually.

### B. CytoTerra library construction and sequencing

Study samples were provided to Phase Genomics as deidentified cryopreserved specimens. Samples were thawed and cell numbers were counted by hemocytometer before storage in Phase Genomics’ proprietary preservative PGShield™. Samples were then stored at 4°C prior to library preparation.

Libraries were prepared from 200,000-500,000 cells using the Phase Genomics CytoTerra kit (v1.1) following the manufacturer’s protocol. In brief, cells suspended in PGShield were crosslinked, quenched, and bound to magnetic recovery beads. Bead-bound cells were then washed and lysed prior to restriction digestion and fill-in with biotinylated nucleotides. Digested chromatin fragments were subjected to proximity ligation. Following proximity ligation, chromatin crosslinks were enzymatically reversed, and proximity ligation products purified using streptavidin beads. Streptavidin bead-bound fragments were then used to generate an Illumina-compatible paired-end sequencing library. Libraries were sequenced on a NovaSeq 6000 in the paired-end 150 bp format to an average depth of 150M read pairs. Library performance was evaluated using Phase Genomics’ open-source quality control tool hic_qc(6). Libraries passing QC were advanced to analysis.

### C. Analysis of structural variants on the CytoTerra platform

Raw FASTQ files for each sample were used as inputs for the CytoTerra cloud-based analysis platform. Reads were mapped to the human genome reference (GRCh38) using BWA using the Hi-C flags(7). Aligned reads were processed by a suite of proprietary bioinformatic and artificial intelligence (AI) variant callers to evaluate the presence of structural variants. Call sets generated by the platform were integrated and presented for review using Curator, the interactive evaluation tool on the CytoTerra cloud platform. All variant calls were evaluated manually prior to reporting. Analysis of GPM data was performed blinded to the known cytogenetic lesions of the AML cases.

## Results

### D. Clinical AML samples performance in proximity ligation sequencing

Cryopreserved AML samples were identified from the FH AML sample repository. The study population is composed of 48 diagnostic samples derived from peripheral blood, bone marrow aspirates, and apheresis samples. Estimates of blast percentage varied between 5% and 96% (Fig. 2A). GPM libraries generated from between 200,000 and 500,000 cells were successful for all samples. Post-sequencing quality control analysis of the resulting libraries showed acceptable performance for all libraries (Fig. 2B).

**Fig. 2.**
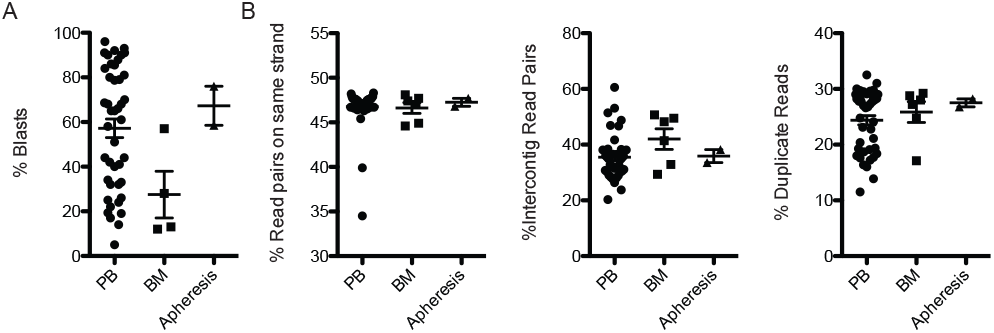
Blast counts and library parameters for samples used in this study. (A) Blast percentage estimates for peripheral blood (PB), bone marrow aspirates (BM), or apheresis-derived samples used for GPM library construction. (B) Quality control metrics for libraries generated by sample type. Reads on same strand measures the percent of read pairs from library inserts that map to the same strand of the human reference genome and are therefore the product of a proximity ligation event. Because reads and be proximity ligated to either the same or different strand configuration, the same strand percentage multiplied by 2 gives an estimate of the fraction of library fragments derived from proximity ligation events. Inter-contig mapping read pairs measures the fraction of read pairs likely to be derived by spurious ligation. Duplicate reads are the fraction of reads that are the result of either PCR or optical duplication.

### E. Detection of inter-chromosomal and intrachromosomal rearrangements

Based on previous cytogenetic risk assessments, The blinded CytoTerra assessment identified all ELN-identified translocations in the study population. These include the t(8;21)(q22;q22.1) that generates the *RUNX1::RUNXT1* fusion (Fig. 3A). Beyond the ELN-classified translocation identified, several additional rearrangements of known and unknown significance were observed in the CytoTerra analysis. These variants included a *NUP98::KDM5A* fusion created by a t(11;12)(p15.4;p13.33), a variant associated with poor prognosis and chemoresistance (Fig. 3D)(8). This rearrangement was not previously detected in the cytogenetics report for this patient. Though not previously reported in the literature, a t(6;7)(p23;q36.3) identified demonstrated a small region of non-reciprocal exchange resulting in a deletion of the *JARID2* gene, a known tumor suppressor in myeloid neoplasms (Fig. 3E)(9). The remaining variants identified were of no known significance (Supplementary File 1). However, based on ELN 2022 risk classification, these variants can contribute to a ‘masked complex karyotype,’ defined as three or more unrelated chromosome abnormalities(2). We observed a single complex karyotype in this study population that included 6 inter-chromosomal breakpoints, in addition to a -7 and dup(21)(q22.12) (Fig. 4C) amongst other less complex karyotype (Fig. 4A-B).

**Fig. 3.**
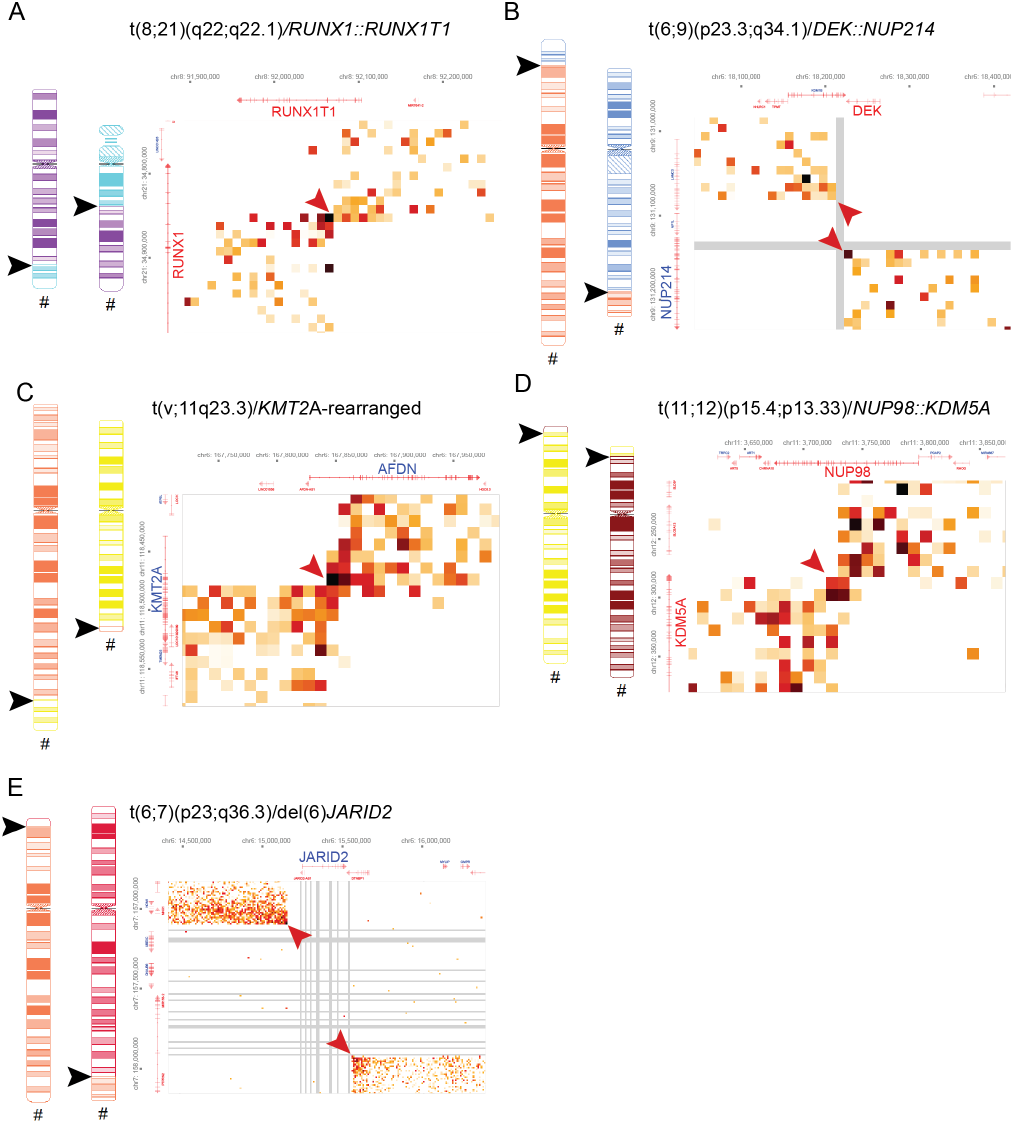
Selected example heatmaps of translocations detected in this study. Arrowheads indicate breakpoints observed on ideograms (black) and heatmaps (red). Coordinates of pair-wise interactions and associated gene models are labelled on X and Y axes. Gray bars indicate a lack of detected pair-wise interaction.

**Fig. 4.**
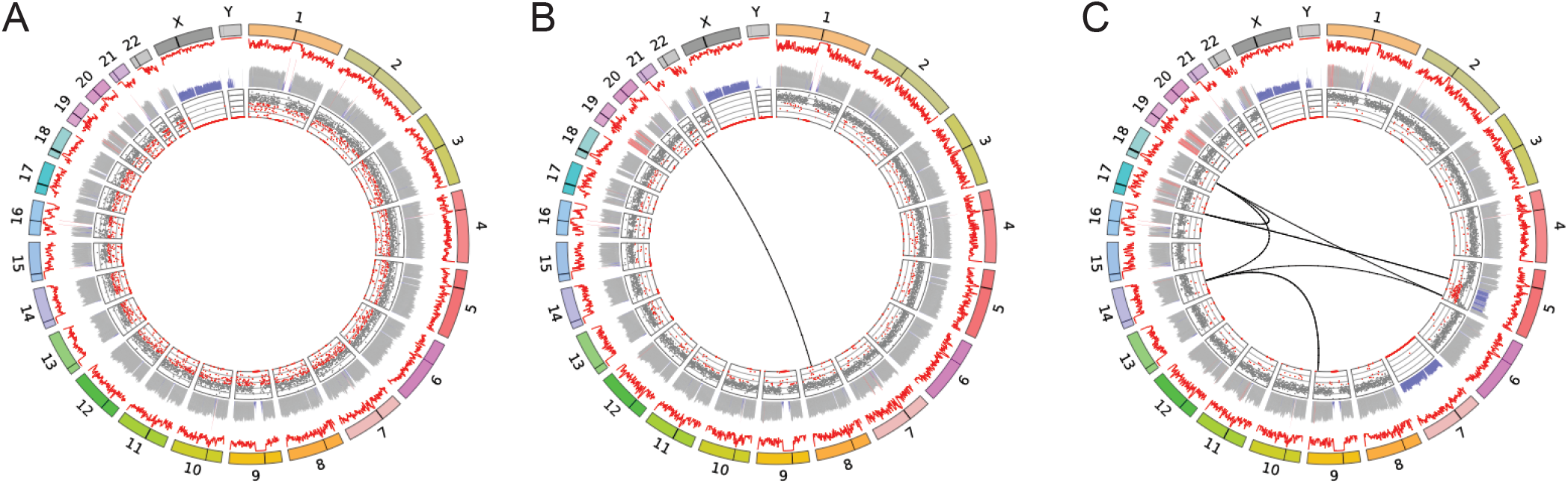
Circos plots illustrating the range of complexity observed in this study. (A) A normal karyotype patient sample (46, XY). (B) A patient presenting with 45,X t(8;21)(q22;q22.1). (C) A complex series of genomic rearrangements uncovered using GPM. Inferred ISCN for this case: 45,XY,del(5)(q22q35),der(5)t(5;17)(q14.3;p13.3),- 7,der(9)t(9;14)(q33.3;q23.1),der(14)del(14)(q22.2q23.1)t(9;14),der(17)t(17;18)(p13.3;q21.1),der(18)(18pter->18q21.1::14q23.1::5q35.1->5qter). Outer ring: Chromosomes represented as colored boxes, black bar illustrates the location of the centromere. Middle rings: Red line illustrates raw coverage with inferred copy number illustrated as bars below. Gray = copy 2, blue = copy 1, red > copy 2. Inner ring: minor allele frequency (MAF), gray dots indicate expected MAF for copy 2 while red dots indicate a deviation from expected frequency.

Among this data set, inv(16)(p13.1q22) is the most common, observed in between 4-5% of AML patients that receive cytogenetic work-ups(10, 11). CytoTerra identified 4/4 inv(16) rearrangements observed by cytogenetics in the study population (Fig. 5A). The next most common class of inversions observed are related rearrangements involving the *MECOM* (*EVI1*) locus(11). In this study, both cytogenetics and CytoTerra identified a single instance of an inv(3)(q21.3q26.1) (Fig. 5B). However, only CytoTerra identified an inv(3)(p24.3q26.2), an unusual but previously documented rearrangement(12). Both of these rearrangements are classified as adverse by the ELN classification(2). We also observed an instance of an inv(12)(p13.32p13.2), a variant that has not been previously documented (data not shown). This variant of unknown significance does disrupt *CCND2*, a gene frequently mutated in core-binding factor-driven AMLs(13) but has an uncertain effect on outcomes. Among the 48 cases in this set, we observed a recurrent in-version not previously documented in AML in two cases. The inv(9)(p13.3p13.1) was seen in two unrelated cases and occurs between two paralogous genes *ANKRD18A* and *ANKRD18B* (Fig. 5C). These genes lie within regions of segmental duplication in the pericentromeric region of chr9 at a distance of approximately 5 Mbp from each other.

**Fig. 5.**
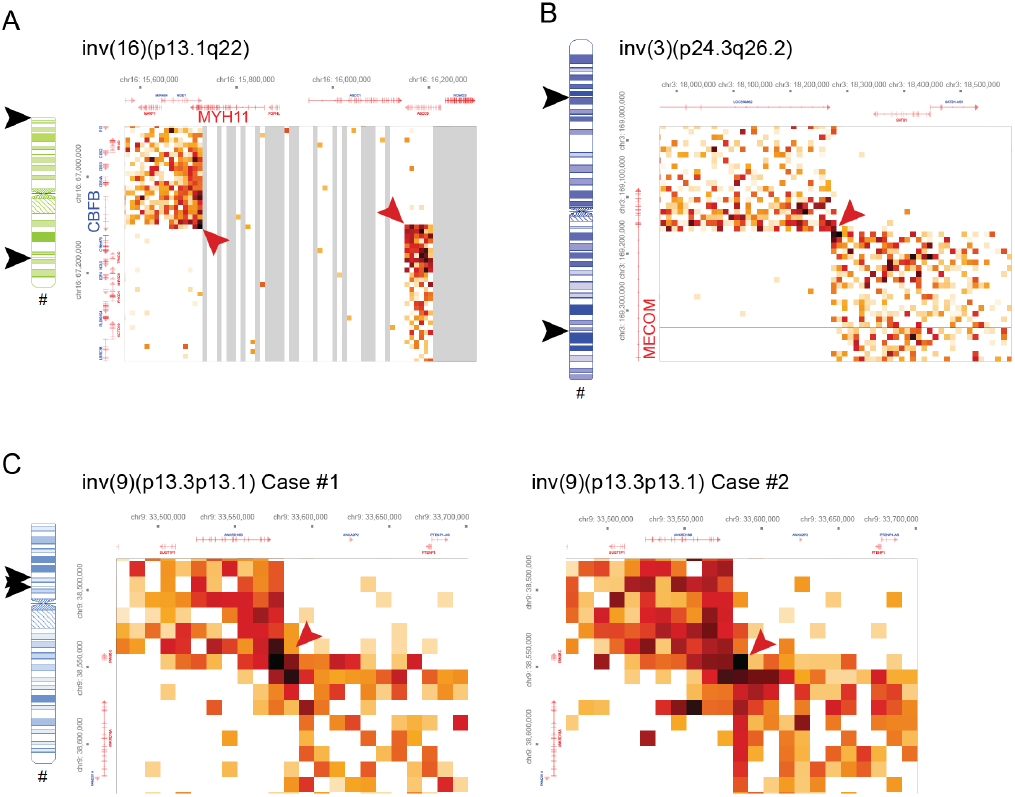
Selected inversions identified in the study population. (A). Example of the most common recurrent inversion observed in AML cases involving chr16, creating a *MYH11::CBFB* fusion gene. (B) A less common inv(3) involving *MECOM* (*EVI1*). (C) A pair of recurrent inv(9) observed in this study, a variant not previously associated with AML. Arrowheads indicate breakpoints observed on ideograms (black) and heatmaps (red).

### F. Detection of insertions

In this case study populations, we identified two insertions, neither of which were reported by cytogenetic analysis. In one case, 8 Mbp of chr5 is inserted into chr13 150 kbp upstream of *FLT3* and disrupting the *PAN3* gene (Fig. 6A). The second insertion of 120 kbp of chr12 into another site on chr12 (Fig. 6B). This small insertion is copy-neutral but does disrupt *DDX11*, a gene that, when mutated, is associated with negative outcomes in AML(14).

**Fig. 6.**
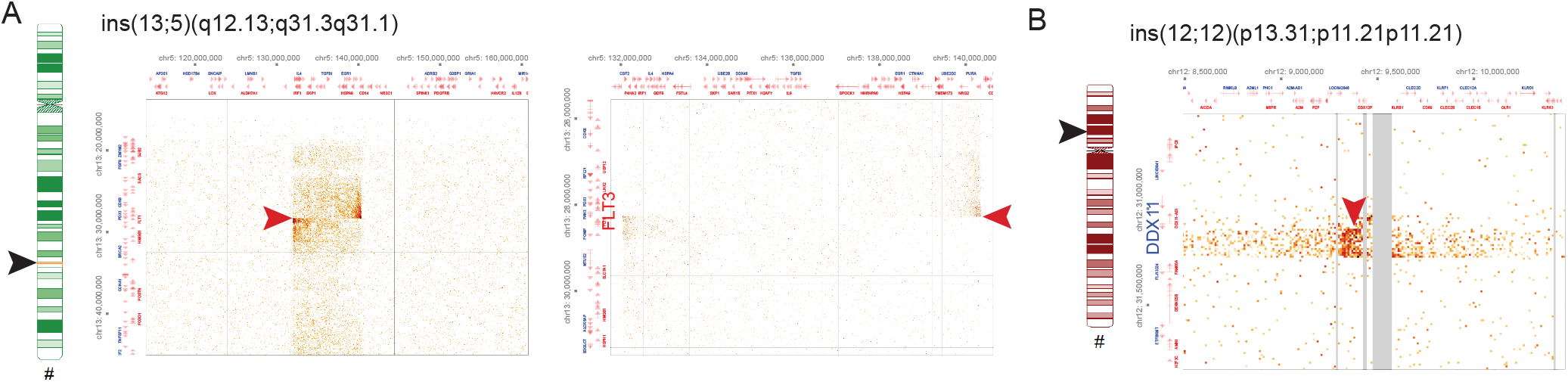
Two insertions observed in the study cohort involving inter- (A) and intrachromosomal(B) DNA sequences. Arrowheads indicate insertion site on ideograms (black) and on heatmaps (red).

### G. Detection of copy number aberrations

In the current study population, only two observations of -7 were detected CytoTerra. Numerous aneuploidies and smaller deletions and duplications of unknown significance were detected by CytoTerra (Supplementary File 1). Amongst these are two incidences of deletions of the *TET2* (Fig. 7A) gene not previously detected in cytogenetic reporting. Of less clear significance, we observed two instances of a recurrent dup(3)(q26.31q26.31) which spans exon 4 of the *NLGN1* gene, a cell surface protein not previously associated with AML.

**Fig. 7.**
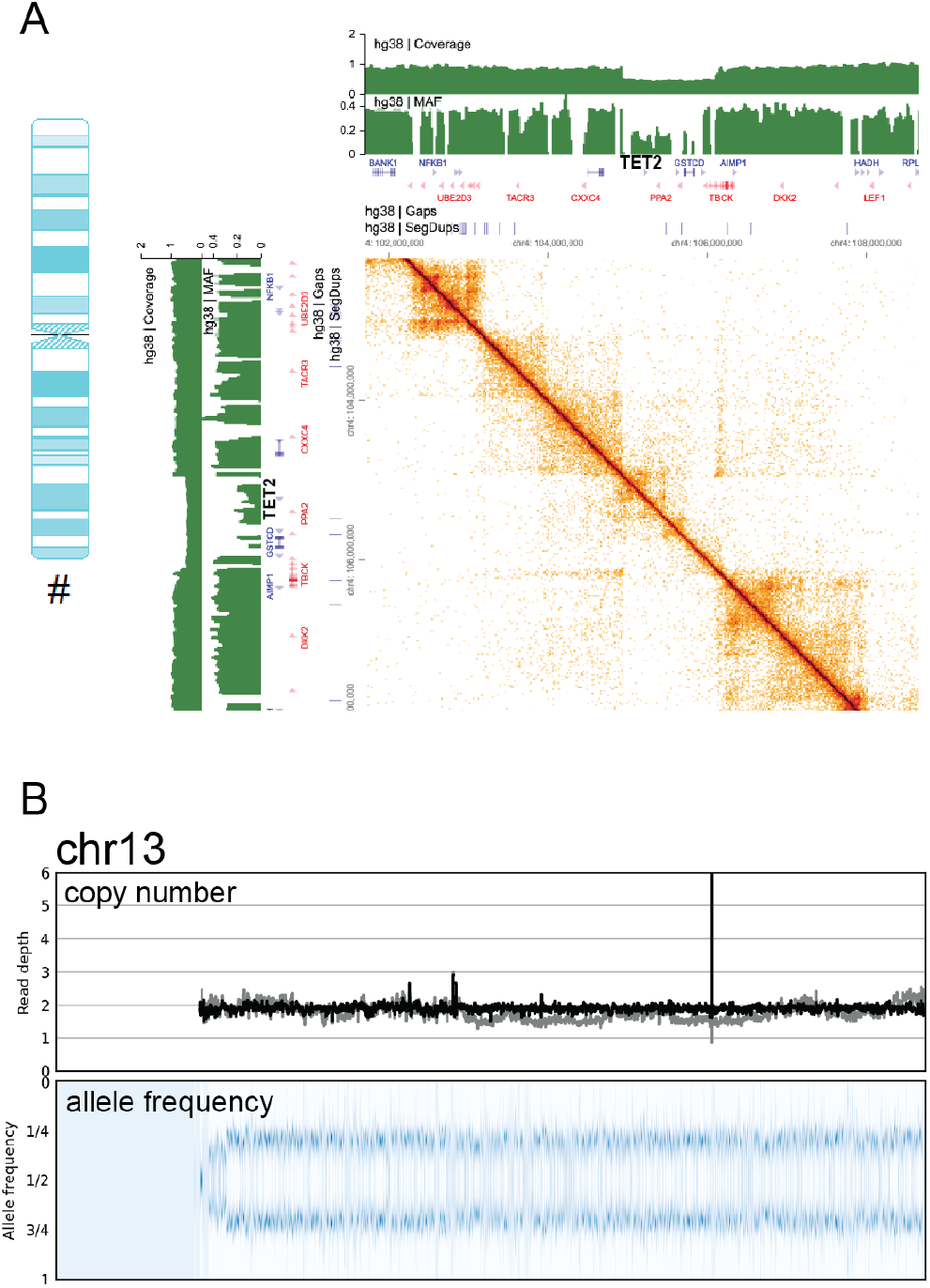
Examples of copy number alteration and copy-neutral loss of heterozygosity (cnLOH) detected by GPM. (A) A del(9) that deletes the CHIP-associated gene *TET2* visualized as a decrease in read coverage (upper green bar plot) and minor allele fraction (MAF, lower green bar plot). Novel contact created by the deletion of intervening sequence are observed on the off-diagonol region of the heatmap. (B) Mosaic cnLOH observed in a case where coverage (upper panel) is copy 2 (gray: raw read depth, black GC-corrected read depth) and a skew in the allele frequency that deviates from 1/2 (Lowe panel).

Copy-neutral loss of heterozygosity (cnLOH), though not associated with ELN Risk categories, is also routinely assessed by CMA. In the study population, two instances of cnLOH of chr13 (Fig. 7B) as well as terminal segments of 6p and 13q were detected by both CMA and CytoTerra.

### H. Concordance between cytogenetics and CytoTerra

Blinded review of variant calls generated by the CytoTerra platform were compared with the record of clinical cytogenetics. CytoTerra showed 100% concordance for all specific variants that have associated impacts on risk stratification as defined by ELN 2022 criteria Table 2. Notably, the percentage of blasts did not have a clear effect on the ability to detect these variants, with % blast ranging between 5 and 96%. When considering all variants reported by cytogenetics, CytoTerra demonstrates a 72.3% concordance rate with cytogenetics (Table 2). A majority (9/13) of discordant calls are aneuploidies (two +8, one +13, one -21, two +22, one -Y and one case of 4N, tetraploidy).

**Table 1.** Clinical demographics

|  |  |
| --- | --- |
| **Total patients** | 48 |
| **Sex** | n(%) |
| Male | 28 (57) |
| Female | 21 (43) |
| **Age, year** | mean (range) |
|  | 54 (21-84) |
| **Specimen Source** | n(%) |
| Peripheral blood | 42 (88) |
| Bone marrow | 4 (8) |
| Apheresis | 2 (4) |

**Table 2.**
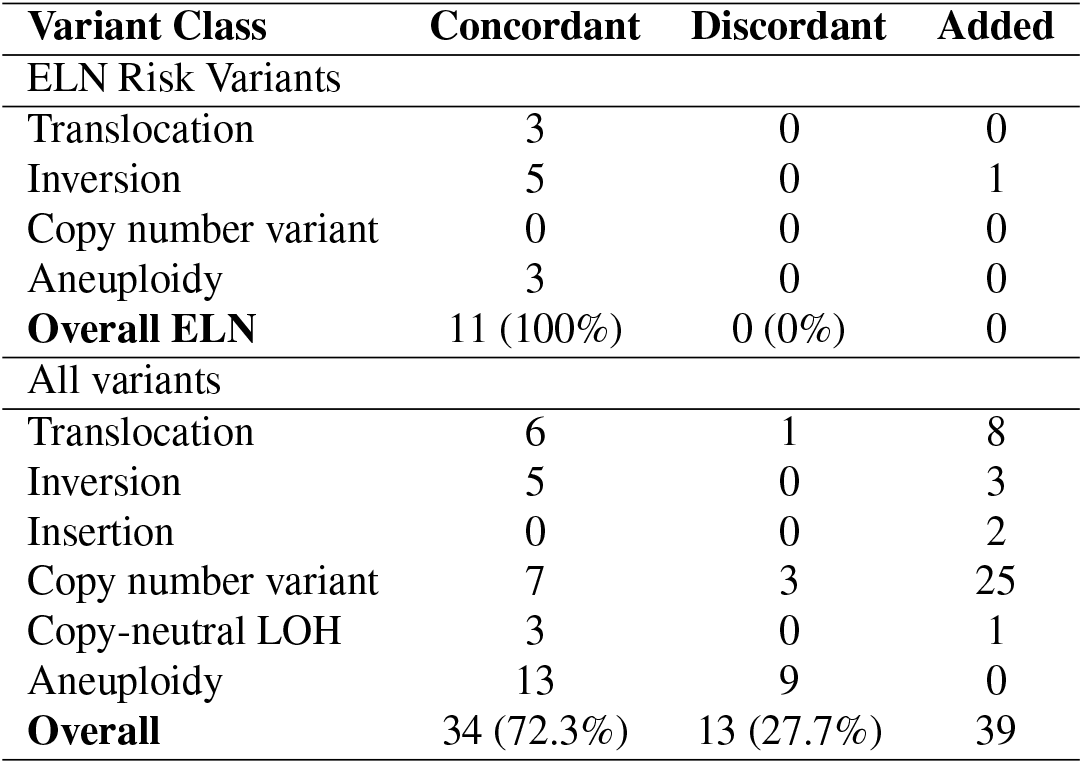
Summary of concordance between PGM and standard-of-care cytogenetics

| Variant Class | Concordant | Discordant | Added |
| --- | --- | --- | --- |
| ELN Risk Variants |  |  |  |
| Translocation | 3 | 0 | 0 |
| Inversion | 5 | 0 | 1 |
| Copy number variant | 0 | 0 | 0 |
| Aneuploidy | 3 | 0 | 0 |
| **Overall ELN** | 11 (100%) | 0 (0%) | 0 |
| All variants |  |  |  |
| Translocation | 6 | 1 | 8 |
| Inversion | 5 | 0 | 3 |
| Insertion | 0 | 0 | 2 |
| Copy number variant | 7 | 3 | 25 |
| Copy-neutral LOH | 3 | 0 | 1 |
| Aneuploidy | 13 | 9 | 0 |
| **Overall** | 34 (72.3%) | 13 (27.7%) | 39 |

## Discussion

### Summary

This study shows the ability of whole genome sequencing with GPM to detect cytogenetic aberrations including rearrangements, inversions, copy number alterations, insertions and deletions, as well as copy neutral loss of heterozygosity. Over an initial set of AML patients representing the full range of ELN prognostic risk categories, we challenged the GPM assay to recapitulate the prognostic data produced by archival clinical grade molecular and cytogenetic assays.

### Impact of GPM on ELN risk stratification

The ELN 2022 risk classification guidelines identifies a set of recurrent translocations seen in AML. Though less commonly observed than translocations, a number of recurrent inversions are known contributors to the AML phenotype. The *RUNX1::RUNXT1* fusion is associated with favorable outcomes and may be an important biomarker for treatment with CDK4/6 inhibitors(15). Inv(16)(p13.1q22) is the most common intrachromosomal rearrangement and generates a *CBFB::MYH11* fusion protein, a rearrangement that carries with it a more favorable prognosis. The t(6;9)(p23.3;q34.1) similarly generates a *DEK::NUP214* fusion gene but is associated with poor prognosis(Fig. 3B) 16). Members of the nucleoporin gene family, including *NUP98* and *NUP214*, are known to drive AML through a variety of different partner genes that are detectible using the GPM approach. Like the nucleoporin genes, *KMT2A* is known to associate with a variety of fusion partners(17), including the AFDN gene observed in this study population (Fig. 3C). While standard-of-care cytogenetics identified both *KMT2A* and *NUP214* fusions, only CytoTerra identified the *NUP98* fusion, a variant associated with poor outcomes(8). In this case, CytoTerra offers benefit in identifying variants of import over existing standard methods.

### GPM discovers novel findings

Detecting insertions by traditional cytogenetics is dependent on the size and genomic content of the inserted DNA segment. Some insertions are routinely detected over the course of AML diagnosis (i.e. *FLT3*-ITD) but most of the variants are anonymous and are of unknown significance. In this study, GPM detected 2 insertions which were not detected Mbp cytogenetics one 8Mbp, and one 120Kbps in length. The resolution of GPM enables more thorough description of the insertions identified in this study, both of which are associated with genes (*FLT3, DDX11*) of known clinical import in AML. Interestingly we observed a recurrent inversion that has not been previous documented in AML genomes in two of our cases involving inv(9)(p13.3p13.1 between two paralogous genes *ANKRD18A* and *ANKRD18B* spanning a 5MB region and lie in a pericentromeric region and involve a sub-microscopic interval of the genome. Rearrangements like this can be challenging to detect by cytogenetics and standard short read sequencing methods. However, this variant has been observed previously in a number of case of acute lymphoblastic leukemia(18), detected only through a targeted resequencing effort of this pericentromeric region. These cytogenetically cryptic classes of rearrangement represent a relevant class of variant where CytoTerra shows particular promise to make a clinical impact.

Unsurprisingly, the majority of additional variants called by CytoTerra involve copy number changes below the level of cytogenetic resolution (<5 Mb). They may also reflect changes deemed unreportable from a clinical perspective as many of these variants are of unknown significance, including potentially constitutional variants. However, as noted above, CytoTerra identified additional variants of known significance not specified in the ELN risk criteria, including *NUP98::KDM5A*. This, and six other translocations, and two additional inversions, were uncovered by CytoTerra in this study. Previous cytogenetic analysis likely failed to observe these rearrangements because of the genomic location and/or size of the genomic interval involved. For example, NUP98 lies at the 11p terminus and has multiple oncogenic partners making it particularly challenging to detect.

### Limitations of GPM

Relatively few copy number aberrations (CNAs) are considered to be informative for risk stratification by the ELN 2022 guidelines. CNAs can also be included under the catch-all category of ‘cytogenetic and/or molecular abnormalities not classified as favorable or adverse’, which impart moderate risk assessment. These include -5, del(5q), -7, -17 (or -17p), all of which are associated with adverse risk. One of the limitations of GPM is its relatively lower sensitivity in detecting copy number alterations. This highlights the challenge of detecting mosaic changes in whole chromosome copy number in all sequencing-based coverage data. Tetraploidy also represents a challenge because, in the case of whole genome duplication, the allele frequency remains in balance and is undetectable by sequencing, array, or optical genome mapping. Though not strictly associated with outcomes, *TET2* mutations are a common feature of clonal hematopoiesis of indeterminate potential (CHIP)(19). In this study although 2 cases demonstrated copy number alterations around *TET2*, it is uncertain if these are associated with the AML or the underlying mutational background attributable to CHIP. A limitation of this study is the relative low number of cases of some specific chromosomal aberrations such as having only one patient with cnLOH, however this was in a case that was confirmed by CGAT. CnLOH of 13q is frequently observed in AML with *FLT3*-ITD mutations(20) and has been reported to be associated with an adverse outcome in AML(21). The limitations described above may be overcome by increased depth of sequencing, allowing for higher confidence detection of subtle changes of minor allele frequency.

In this study we have demonstrated GPM’s capability to comprehensively interrogate the entire genome including detecting cryptic chromosomal aberrations at a higher resolution than conventional karyotyping and chromosomal genomic array testing. The identification of a novel recurrent AML variant in this 48-sample study demonstrates the potential of GPM as a tool for biomarker discovery. The improved detection of ELN risk variants with GPM warrants a comprehensive study to evaluate CytoTerra for improved accuracy in patient risk stratification.

## Data Availability

Data will be made publicly available through dbGAP upon acceptance at a peer-reviewed journal.

## Acknowledgments

This work was supported by an SBIR Phase II grant from NCI/NIH R44CA278140 to SME and Phase Genomics. This work is also partially supported by UG1 CA233338-02, and CA175008-06 to JR at FHCC.

## Author contributions

For every author, his or her contribution to the manuscript needs to be provided using the following categories: Conception: CCSY, SME, IL, MF, JR Interpretation or analysis of data: CCSY, SME, MM, MF, JR Preparation of the manuscript: CCSY, SME Revision for important intellectual content: MF, JR Supervision: CCSY, SME

## Data Availability

This is an active project with ongoing enrollment of subjects for the study thus the data will be made publicly available at a later date when the study is ended. However, data presented in this study will be made available upon request to the corresponding author, please contact.

## Bibliography

1. T.S.K. Wan. Cancer cytogenetics: An introduction. In T.S.K. Wan, editor, Cancer Cytogenetics, pages 1–10. Springer New York, New York, NY, 2017.

2. H. Döhner, A.H. Wei, F.R. Appelbaum, C. Craddock, C.D. DiNardo, et al. Diagnosis and management of aml in adults: 2022 recommendations from an international expert panel on behalf of the eln. Blood, 140:1345–1377, 2022.

3. J.N. Burton, A. Adey, R.P. Patwardhan, R. Qiu, J.O. Kitzman, and J. Shendure. Chromosome-scale scaffolding of de novo genome assemblies based on chromatin interactions. Nature Biotechnology, 31:1119–1125, 2013.

4. N. Kaplan and J. Dekker. High-throughput genome scaffolding from in vivo dna interaction frequency. Nature Biotechnology, 31:1143–1147, 2013.

5. E. Lieberman-Aiden, N.L. Van Berkum, L. Williams, et al. Comprehensive mapping of longrange interactions reveals folding principles of the human genome. Science, 326:289–293, 2009.

6. Phase Genomics. Hic_qc, n.d. https://github.com/phasegenomics/hic_qc.

7. H. Li and R. Durbin. Fast and accurate short read alignment with burrows-wheeler transform. Bioinformatics, 25:1754–1760, 2009.

8. S. Mohanty. Nup98 rearrangements in aml: Molecular mechanisms and clinical implications. Onco, 3:147–164, 2023.

9. H. Celik, W.K. Koh, A.C. Kramer, et al. Jarid2 functions as a tumor suppressor in myeloid neoplasms by repressing self-renewal in hematopoietic progenitor cells. Cancer Cell, 34: 741–756.e8, 2018.

10. D. Grimwade, H. Walker, F. Oliver, et al. The importance of diagnostic cytogenetics on outcome in aml: Analysis of 1,612 patients entered into the mrc aml 10 trial. Blood, 92: 2322–2333, 1998.

11. M. Bendari, N. Khoubila, S. Cherkaoui, et al. Current cytogenetic abnormalities in acute myeloid leukemia. In T. Aşkin Çelik and S. Dey, editors, Chromosomal Abnormalities. IntechOpen, 2020.

12. C. Haferlach, U. Bacher, V. Grossmann, et al. Three novel cytogenetically cryptic evi1 rearrangements associated with increased evi1 expression and poor prognosis identified in 27 acute myeloid leukemia cases. Genes Chromosomes & Cancer, 51:1079–1085, 2012.

13. A.-K. Eisfeld, J. Kohlschmidt, S. Schwind, et al. Ccnd1 and ccnd2 mutations are frequent in adults with core-binding factor acute myeloid leukemia (cbf-aml) with t(8;21)(q22;q22). Blood, 128:2740–2740, 2016.

14. T. Zhang, P. Auer, J. Dong, et al. Whole-genome sequencing identifies novel predictors for hematopoietic cell transplant outcomes for patients with myelodysplastic syndrome: a cibmtr study. Journal of Hematology & Oncology, 16:37, 2023.

15. L.E. Swart and O. Heidenreich. The runx1/runx1t1 network: translating insights into therapeutic options. Experimental Hematology, 94:1–10, 2021.

16. M.L. Slovak, H. Gundacker, C.D. Bloomfield, et al. A retrospective study of 69 patients with t(6;9)(p23;q34) aml emphasizes the need for a prospective, multicenter initiative for rare ‘poor prognosis’ myeloid malignancies. Leukemia, 20:1295–1297, 2006.

17. M. Bill, K. Mrózek, J. Kohlschmidt, et al. Mutational landscape and clinical outcome of patients with de novo acute myeloid leukemia and rearrangements involving 11q23/ kmt2a. Proceedings of the National Academy of Sciences, 117:26340–26346, 2020.

18. V.K. Sarhadi, L. Lahti, I. Scheinin, et al. Targeted resequencing of 9p in acute lymphoblastic leukemia yields concordant results with array cgh and reveals novel genomic alterations. Genomics, 102:182–188, 2013.

19. S. Asada and T. Kitamura. Clonal hematopoiesis and associated diseases: A review of recent findings. Cancer Science, 112:3962–3971, 2021.

20. D.L. Stirewalt, E.L. Pogosova-Agadjanyan, K. Tsuchiya, et al. Copy-neutral loss of heterozygosity is prevalent and a late event in the pathogenesis of flt3/itd aml. Blood Cancer Journal, 4:e208–e208, 2014.

21. Christine M. Gronseth, Scott E. McElhone, Barry E. Storer, et al. Prognostic significance of acquired copy-neutral loss of heterozygosity in acute myeloid leukemia. Cancer, 121(17): 2900–2908, 2015.

